# Shift work and evening chronotype are related to prevalent non-alcoholic fatty liver disease in 282,303 UK biobank participants

**DOI:** 10.1101/2022.05.19.22275307

**Authors:** Robert Maidstone, Martin K. Rutter, Thomas Marjot, David W. Ray, Matthew Baxter

## Abstract

**Background & Aims:** Non-alcoholic fatty liver disease (NAFLD) is globally prevalent and confers a high risk of morbidity via progression to non-alcoholic steatohepatitis (NASH). Circadian disruption in mouse models contributes to the development of hepatic steatosis and inflammation, however evidence in humans is lacking. We investigated how shift working and chronotype were associated with NAFLD/NASH in UK Biobank participants.

**Methods:** We stratified 282,303 UK Biobank participants into day, irregular-shift, and permanent night-shift workers. We compared the likelihood of NAFLD/NASH in these groups using: a) Dallas Steatosis Index (DSI), b) NAFLD/NASH ICD10 codes, and c) liver proton density fat fraction (PDFF) after serially adjusting for age, sex, ethnicity, sleep, alcohol, smoking, and body mass index. We further assessed the relationship of baseline chronotype with likelihood of NAFLD/NASH using the same outcomes and covariates.

**Results:** Compared to day workers, irregular-shift workers were more likely to have NAFLD/NASH defined by high DSI (odds ratio (OR) 1.29 (95% CI 1.18–1.4)) after adjusting for all covariates excluding BMI, with some attenuation after additional adjustment for BMI (OR 1.12 (1.03-1.22)). Likelihood of DSI-defined NAFLD/NASH was also higher in permanent night-shift workers (OR 1.08 (0.9–1.29)) in the fully-adjusted model. Compared to participants with intermediate chronotype, those with extreme late chronotype had a higher likelihood of DSI-defined NAFLD/NASH (OR 1.45 (1.34–1.56)) and a higher likelihood of NAFLD/NASH by ICD10 code (OR 1.23 (1.09–1.39)). Liver PDFF was elevated in irregular shift workers, but not permanent night shift workers.

**Conclusions:** Irregular-shift work and chronotype are associated with NAFLD/NASH, suggesting circadian misalignment as an underlying mechanism. These findings have implications for health interventions to mitigate the detrimental effect of shift work.

## Introduction

NAFLD affects over 25% of the worldwide adult population, and the prevalence is increasing [1]. Furthermore, NAFLD is thought to be hugely under-diagnosed clinically. NAFLD is defined by the presence of steatosis in >5% of hepatocytes without a secondary cause such as significant alcohol consumption [2]. Approximately 10-30% of people with NAFLD develop non-alcoholic steatohepatitis (NASH), which is associated with progression to liver fibrosis, cirrhosis, hepatocellular carcinoma and cardiovascular disease [1]. It is therefore of critical importance to establish the causes of this global epidemic of NAFLD/NASH.

Shift-working is increasingly prevalent globally. It imposes strain on the circadian timing system, which synchronizes physiology to the external environment, following cues from sunlight, and eating schedules. The core circadian clock is sited in the hypothalamic suprachiasmatic nucleus, where it receives light information from the retina. The time-keeping mechanism is composed of a transcriptional-translational feedback loop, which has an intrinsic period of approximately 24 hours. Circadian rhythms are critically important for healthy regulation of metabolism and endocrine functions [3]. Environmental, and behavioural changes such as artificial light, jet-lag, and shift-work drive misalignment between the circadian clock phase and the external environment [4]. The existing literature is conflicting regarding the relationship between shift-work and NAFLD. A study of 6881 Chinese steel-workers found that rotating night shift work was associated with a higher likelihood of NAFLD as defined by ultrasound, with risks increasing with longer duration of shift-work and longer night-time hours [5]. Another study, incorporating 4740 male workers, found an association of night shift-work with elevated alanine transaminase (ALT) levels [6]. However, the largest prior study included 8159 workers and showed no association between shift-work and NAFLD as defined by liver enzyme levels [7]. All of these studies have significant limitations, including relatively modest sample sizes, lack of comparison between rotating and permanent night shift-workers, and use of imprecise methods for diagnosing NAFLD. Studies were also restricted to single occupations and racial groups which limit external generalizability.

Here, we aim to address the limitations of prior work by providing a comprehensive analysis of associations between several shift-work sub-types and NAFLD/NASH using a large and diverse UK cohort in which a range of methods for diagnosing NAFLD/NASH are available. Further, we test the potential mechanistic role of circadian disruption in NAFLD/NASH by examining the relationships with chronotype, the intrinsic preference for morning or evening activity. Individuals with evening chronotype are known to have a greater likelihood of experiencing circadian disruption because their natural sleeping patterns are more likely to be disturbed by regular work routines than morning chronotypes. We find that NAFLD/NASH is associated with certain shift-work schedules, particularly those who work irregular night shifts. NAFLD/NASH is also significantly associated with extreme evening chronotype, indicating a plausible pathogenic role for circadian misalignment: conflict between the internal clock time, and environment (light-dark), and behaviour (eating pattern).

## Methods

The UK Biobank recruited 502,540 participants, registered with the UK National Health Service (NHS) between 2007-2010. Participants were aged 40-69 years and completed baseline questionnaires on occupation, work hours, medical history and lifestyle. Further information on medical conditions, medication and health status was gathered by trained health professionals.

### Assessment of shift-work

Analysis of shift work was performed on participants who were in paid employment or self-employed at time of baseline assessment (n =286,825; Supplementary Table 1). Participants were asked the following questions: Does your work involve shift work? The options presented were: Never/Rarely, Sometimes, Usually, Always, Do not know, Prefer not to answer. Those who answered “Do not know” or “Prefer not to answer” were excluded from further analysis. Those who answered “Never/Rarely” were categorized as “Day workers”, and those who answered “Sometimes”, “Usually”, or “Always” were subsequently asked: Does your work involve night shifts? The options presented were: Never/Rarely, Sometimes, Usually, Always, Do not know, Prefer not to answer. Those who answered “Do not know” or “Prefer not to answer” were excluded. Those who answered “Never/Rarely”, “Sometimes”, or “Usually” were categorized as “Irregular shift workers” and those who answered “Always” were categorized as “Permanent night workers”. Participants were thus categorized into 3 distinct groups: day workers (control), irregular shift workers, and permanent night workers

### Diagnosis of NAFLD/NASH

We defined NAFLD and/or NASH using three methods: First, we used the Dallas Steatosis Index (DSI) [8] to define the probability of NAFLD at baseline, which is based on a logit prediction model incorporating age, diabetes, hypertension, triglycerides and ethnicity variables (Appendix A, Supplemental material). Second, we used prior hospital admissions associated with baseline ICD10 codes for either NAFLD and/or NASH. Finally, in an exploratory analysis, we used proton density fat fraction (PDFF) to define liver fat percentage in a sub-group of 6,419 UK Biobank participants undergoing liver MR scanning (between August 2014 and October 2015). We used a 5.5% PDFF threshold to define NAFLD [9], and using this measure compared NAFLD prevalence at follow-up by baseline shiftwork status.

### Definition of Chronotype

Chronotype was self-reported at baseline by answering a questionnaire. Participants were asked whether they consider themselves to be: “Definitely a ‘morning’ person”, or “More a ‘morning’ than an ‘evening’ person”, or “More an ‘evening’ than a ‘morning’ person”, or “Definitely an ‘evening’ person”, or “Don’t know”, or “Prefer not to answer”. We combined people responding, “More a ‘morning’ than an ‘evening’ person” and “More an ‘evening’ than a ‘morning’ person” to form referent group against which we compared outcomes in subjects responding, “Definitely a ‘morning’ person” or “Definitely an ‘evening’ person”. Participants answering “Do not know” or “Prefer not to answer” were excluded. Self-reported chronotype defined in this way has been shown to correlate with dim-light melatonin onset and sleep timing [10–12].

### Statistical Analysis

Multivariate logistic regression models compared adjusted odds ratios and corresponding 95% asymptotic confidence intervals for NAFLD outcomes in different exposure groups, with day workers and people with intermediate chronotype serving as referent groups. Model 1 was adjusted for sex, age, ethnicity and Townsend deprivation index (TDI). Model 2 adjusted for model 1 covariates plus sleep duration (categorized as short sleepers; <6hours, normal sleepers; and long sleepers, >8hours) alcohol intake frequency (“daily or almost daily”, “three or four times a week”, “once or twice a week”, “one to three times a month”, “special occasions only” or “never”), smoking status (current, previous or never), smoking pack years (number of cigarettes smoked per day, divided by twenty, multiplied by the number of years of smoking), and length of working week. Finally, model 3 contained model 2 covariates plus BMI, which could be a mediator and/or potential confounder for the association between shift work and NAFLD/NASH. Participants with missing data for covariates were excluded.

## Results

Out of 286,825 UK Biobank participants studied, 42,786 were irregular shift workers and 7142 were permanent night shift workers, the remainder being day workers (Supplementary Table 1). Irregular and permanent nightshift workers were older, more likely to be male and of non-White ethnicity and had higher BMI values compared to day workers. They were also more likely to smoke but less likely to drink alcohol compared to day workers.

### Irregular and permanent night shift work is associated with greater predicted probability of NAFLD

We selected the Dallas Steatosis Index (DSI) to assess the prevalence of NAFLD, which was recently shown to perform with high accuracy in two large population datasets, including the UK Biobank [8]. We analysed DSI score as a continuous variable in different work schedule populations (Figure 1A, Supp Table 2b), and secondly as a categorical variable using a score >0.6% as a cut-off for the definition of NAFLD (Supp Table 2a).

**Figure 1.**
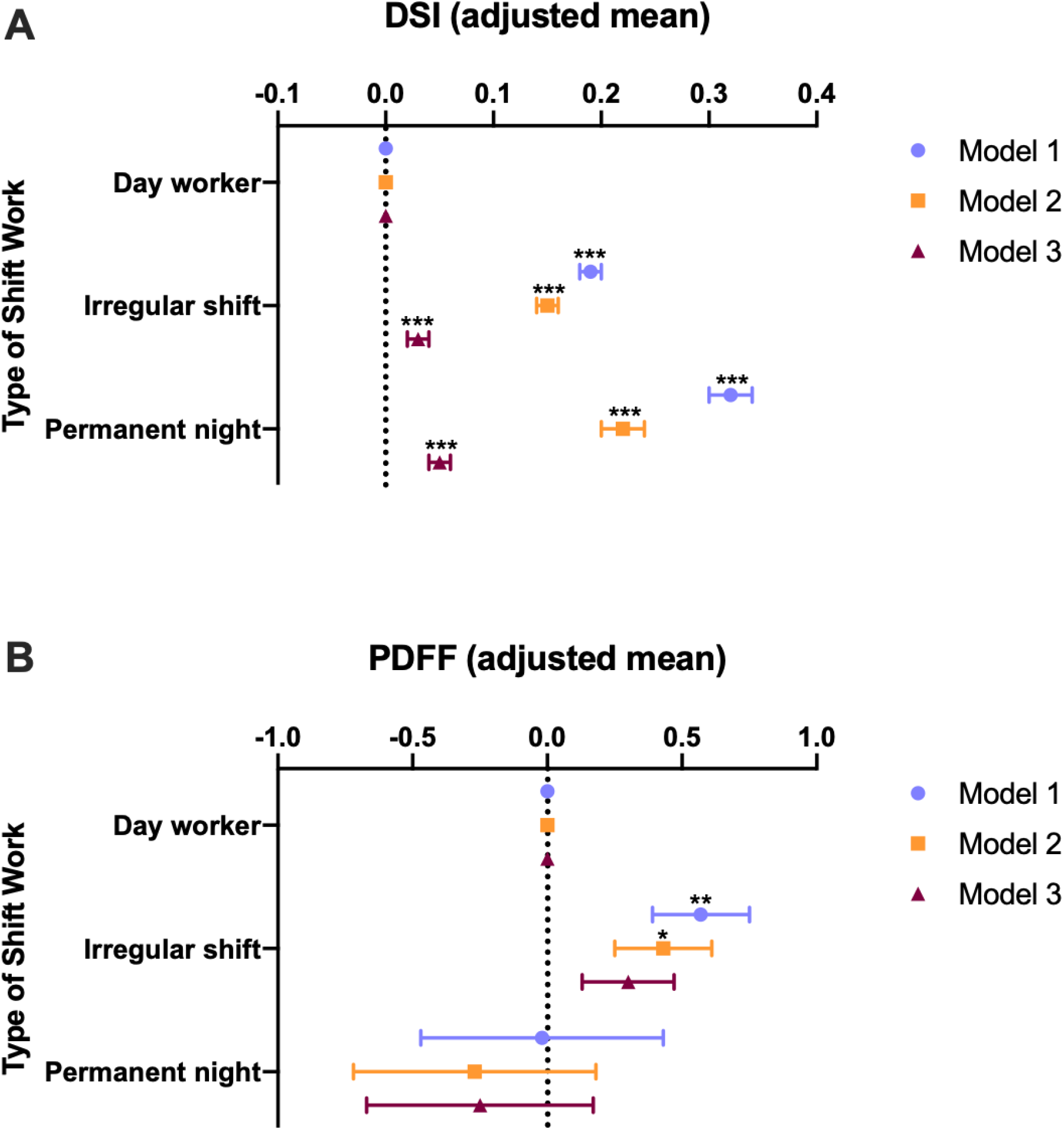
Shift work is associated with NAFLD. Participants were categorized as Day workers, Irregular shift workers, or permanent night shift workers. A. Adjusted mean (95% CI) of Dallas Steatosis Index (DSI), n=240,589. B. Adjusted mean (95% CI) of liver fat as measured by proton density fat fraction (PDFF), n=6,419. Model 1 was adjusted for sex, age, ethnicity and Townsend deprivation index (TDI). Model 2 adjusted for model 1 covariates plus sleep duration alcohol intake frequency, smoking status, smoking pack years, and length of working week. Model 3 contained model 2 covariates plus BMI. Statistical significance was determined by Student’s t-test. * p<0.05; ** p<0.01; ***p<0.001.

Out of 240,589 UK Biobank participants, mean DSI scores were significantly higher in both irregular shift workers and in permanent night shift workers compared to day workers in all 3 models (Figure 1A, Supp Table 2b). Differences in DSI values when comparing shift workers with day workers were markedly attenuated, but remained significant, after adjusting for BMI (model 3; Figure 1A, Supp Table 2b).

When defining risk of NAFLD using DSI score (>0.6), higher odds of NAFLD was observed in the permanent night shift workers compared to day workers in models 1 and 2, but not after further adjustment for BMI in model 3 (Supp Table 2a). Using this method, we found a higher likelihood of NAFLD in irregular shift workers in all 3 models.

### NAFLD/NASH is not significantly linked to NAFLD/NASH by ICD10 code but this may be explained by under-diagnosis

We did not identify a higher likelihood of NAFLD/NASH in shift workers when the diagnosis was based on baseline ICD 10 coding (Supp Table 3). However, the odds ratios had wide confidence intervals suggesting that the analysis was underpowered.

It is recognized that most NAFLD/NASH remains undiagnosed and is missed by ICD10 coding. We observed this in the UK Biobank, where in day workers, irregular shift workers, and permanent night workers, the prevalence of NAFLD or NASH by ICD10 diagnosis of was 0.08, 0.1, and 0.11% respectively (Supp Table 3). NAFLD is thought to have a prevalence >25% in UK. ICD10 code diagnosis is therefore an inappropriate measure to analyse the risk of NAFLD or NASH.

### Irregular shift-work is associated with high liver fat

We next conducted an exploratory analysis of the association between shift work and liver fat, as measured by proton density fat fraction (PDFF). We analysed liver fat as a continuous variable in different work schedule populations (Figure 1B, Supp Table 4B), and secondly as a categorical variable using >5.5% fat as a cut-off for the definition of non-alcoholic fatty liver (Supp Table 4A) [13]. When compared to day shift workers, irregular shift workers, but not permanent night shift workers, had a higher mean PDFF (Figure 1B, model 1) values and a higher odds of elevated liver fat (>5.5%) (Supp Table 4a; model 1). Attenuation of these relationships were observed in models 2 & 3 when the elevated liver fat outcome was considered as a binary outcome variable. However, when PDFF was considered as a continuous variable, irregular shift workers had higher levels of liver fat when compared to day workers in models 1 and 2, although statistical significance was lost after further adjustment for BMI in model 3 (Figure 1B).

### Extreme chronotype is associated with greater risk of NALFD/NASH

We hypothesised that the increased risk of NAFLD/NASH in certain groups of shift-workers may be due to circadian misalignment. In order to test this hypothesis we examined the relationship between chronotype and NAFLD/NASH. Individuals with extreme chronotypes may experience circadian misalignment in a similar way to shift-workers [14,15]. Compared to individuals with an intermediate chronotype, the mean DSI values were significantly higher in the strong eveningness group in all 3 models, although adjustment for BMI (model 3) attenuated the relationship (Figure 2A; Suppl Table 5a). Mean DSI values were also higher in the strong morningness group in models 1 and 2, although this was entirely abrogated in model 3. There was a significant association between strong eveningness and having a high probability of NAFLD (DSI>0.6) and this persisted in all models (Supp. Tables 5a and 5b).

**Figure 2.**
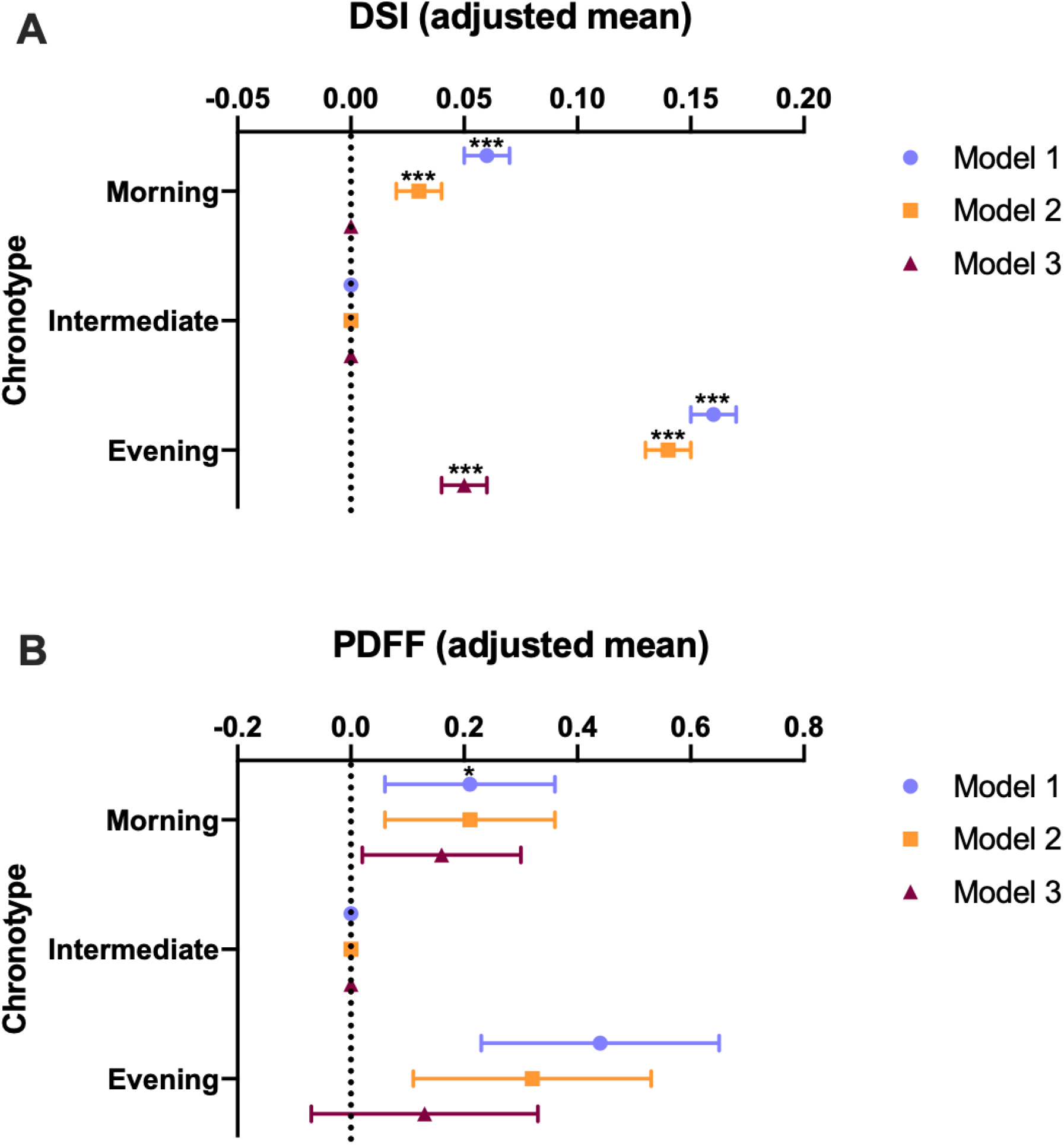
Chronotype is associated with NAFLD. Participants were categorized as extreme morningness (“morning”), extreme eveningness (“evening”), and compared to control “intermediate” chronotype. A. Adjusted mean of Dallas Steatosis Index (DSI), n=377,560. B. Adjusted mean of liver fat as measured by proton density fat fraction (PDFF), n=8,651. Model 1 was adjusted for sex, age, ethnicity and Townsend deprivation index (TDI). Model 2 adjusted for model 1 covariates plus sleep duration alcohol intake frequency, smoking status, smoking pack years, and length of working week. Model 3 contained model 2 covariates plus BMI. Statistical significance was determined by Student’s t-test. * p<0.05; ** p<0.01; ***p<0.001.

In the exploratory analysis, neither strong eveningness, nor strong morningness was significantly associated with high PDFF (>5.5%; Supp. Table 6a). However, average PDFF was higher in the strong eveningness group than the intermediate group in the minimally adjusted model (model 1; Figure 2B).

To examine whether the association between shift work and liver disease was influenced by chronotype, we stratified the participants into three subsets; definite morning, definite evening and intermediate chronotypes. In all chronotype subgroups there was a significant association between shift work and DSI. However, a significant interaction by chronotype status (p_interaction_<0.01), suggested that the relationship between shift work and DSI was stronger in morning than in evening chronotypes (Supplemental Table 8).

## Discussion

In this study we show that shift-workers have higher odds of NAFLD/NASH than non-shift workers. The way in which these relationships were attenuated on adjustment for BMI are in keeping with a mediating role of obesity, which was more prevalent in shift workers when compared to day workers. Furthermore, extreme late chronotype was also associated with increased odds of developing NAFLD/NASH. Individuals with an extreme late chronotype exist in a state of circadian misalignment between their internal circadian clock, and the environmental light-dark cycle, and behavioural activity-feeding-rest cycle. We also find that there is an interaction between chronotype, and shiftwork exposure when it comes to NAFLD/NASH, suggesting that those with morning chronotype may be at greatest risk of developing metabolic liver disease as a consequence of shift work. Taken together, these observations provide evidence that circadian misalignment resulting from shiftwork exposure may contribute to development of NAFLD.

Our findings have immediate and clear clinical and public health implications. NAFLD is an increasingly important global health burden. Cases of NAFLD worldwide have risen from 391.2 million in 1990 to 882.1 million in 2017 [16]. Recent modelling data has forecast that prevalent NAFLD cases will increase 21% between 2015 and 2030, and prevalent NASH cases will increase by 63%. In real terms this projection means a NAFLD prevalence in the adult population of 33.5% by 2030. In the same period, the proportion of NAFLD cases classified as NASH will increase from 20% to 27%. Liver deaths worldwide will increase by 178% [17]. These stark figures underline a burgeoning health crisis, and a worldwide unmet clinical need. Much focus has been drawn to the expansion of “western lifestyles” in terms of diet and behavior as an explanation for this epidemic. In particular, the increasing prevalence of circadian disruptive work patterns and social jet-lag, caused by shift-work, parallels the increasing prevalence of NAFLD and NASH. To date, most research into circadian disruption and the development of NAFLD/NASH has employed mouse models [18,19], and evidence in humans has been conflicting and limited by small sample sizes [5–7]. Here, we present the first evidence confidently associating disrupted circadian schedules to NAFLD and NASH in humans.

Our findings contradict those of Balakrishnan *et al*, and broadly align with those of Zhang *et al* [5,7]. Our study has the clear advantage of scale, allowing us to identify associations with far greater confidence, and to consider a range of possible mediators and confounders. Given our large cohort size we were also able to stratify shift-workers by whether they work irregular shift patterns, or whether they work permanent nights. Interestingly, Zhang *et al* found a positive association between the duration of night shifts and NAFLD as well as with the cumulative length of night shifts and NAFLD. We considered three possible approaches to detect NAFLD cases, of these the DSI had the advantage that all the data was collected at the time of enrolment, when work schedule was captured, and the data was available on a very large number of participants. DSI has also been a well validated predictor of NAFLD including in the UK biobank population[8]. We found an association between both irregular, and permanent night shift work, and the mean DSI score. Interestingly, the strength of relationships were markedly reduced when BMI was included in the models as a covariate. These data are consistent with BMI being a confounder and/or a mediator of the relationship between shift work and NAFLD; our prior work has shown that obesity is associated with selection into shift work [20] and other work suggests that obesity may be a consequence of shift work [21].

It is possible that permanent nightshifts may allow closer alignment of the internal circadian clock phase with the external environmental, and the behavioural cycles. This finding led us to test the role of circadian misalignment in a new analysis on extreme chronotypes, and indeed we discovered an association with extreme late chronotypes, when using the DSI as the diagnostic criterion. This is a striking finding, which led us to test for an interaction between night shift work, chronotype and DSI, which suggested that people with morning chronotypes may be at especially high risk of developing NASH in association with shift work. This intriguing hypothesis will need to be confirmed in prospective studies. Establishing an interaction between these three parameters could have important implications for health screening of shift-workers, and could point to strategies to reduce the risk associated with nightshifts, such as recommended eating schedules. Interestingly, in earlier work we also discovered a relationship between early chronotypes and risk of asthma liked to shift work [14].

Our results should be interpreted within the context of several study limitations. PDFF measurements were only available in a small proportion of the participants (6,482 workers), which resulted in large confidence intervals. Furthermore, the PDFF measurements were not conducted at the same time as work pattern assessments and did not account for differential deaths rates by shift work status during follow-up. We therefore used DSI in order to predict NAFLD using non-direct measures, and did not see the same abrogation of risk in permanent night shift workers. We were also unable to examine the role of shift work in the progression of NAFLD to NASH.

Future studies should aim to identify the role of circadian disruption in the transition from NAFLD to NASH. It is increasingly appreciated that NAFLD and NASH are in fact multi-organ diseases, and studies have uncovered the roles of dysregulation in pancreas, adipose tissue, the immune system and the intestine [22–26]. Each of these systems possesses its own peripheral circadian clock, controlling multiple functions. Synchronisation between the clocks of these various tissues is likely to be crucial to maintaining health function, and desynchrony maybe be a crucial mechanism in tissue dysfunction. Indeed, the immune system is under strict control by the circadian clock, and several inflammatory diseases show strong circadian rhythms in symptoms and pathology, including rheumatoid arthritis, asthma and myocardial infarction [27].

Understanding the role of circadian biology in the development of NALFD/NASH may be critical to developing effective pharmacological and non-pharmacological interventions. Furthermore, the circadian timing of pharmacological interventions may be crucial to improving the efficacy and reducing the off-target effects of candidate drugs [28]. For example, mouse studies have shown how time of day administration of glucocorticoids can avoid undesirable side effects, due to circadian modulation of glucocorticoid receptor cistromes [29]. This approach, known as chronomedicine, is potentially of particular importance in the liver where circadian circuits control lipid metabolism, and xenobiotic detoxification.

## Conclusion

This study shows that shift workers have a higher likelihood of having NAFLD and NASH which may be due to circadian misalignment. Our modelling data are in keeping with obesity having a mediating role in the development of shift work-related NAFLD and interaction effects suggest that individuals with morning chronotype may be at greatest risk. Understanding the mechanisms linking shift work to NAFLD and NASH may identify therapeutic targets or modifying behaviors for shift workers to reduce the disease risk.

## Data Availability

All data produced in the present work are contained in the manuscript

## Supplementary Information

**Supplementary Table 1:** Participant characteristics by shift work status (N = 286,825)

|  | Current work schedule |  |  |
| --- | --- | --- | --- |
|  | Day workers | Irregular shift work | Permanent night shift work |
| N | 236897 | 42786 | 7142 |
| Age (years) | 53.45 (7.11) | 52.46 (7) | 52 (6.86) |
| Sex (% male) | 46.58 | 53.87 | 61.43 |
| BMI (kg/m<sup>2</sup>) | 27.09 (4.65) | 27.97 (4.96) | 28.51 (4.88) |
| **Smoking (%):** |  |  |  |
| Never | 58.1 | 53.3 | 51.99 |
| Previous | 31.91 | 31.43 | 30.03 |
| Current | 9.75 | 14.86 | 17.67 |
| Smoking pack-years | 5.42 (12.21) | 7.55 (14.84) | 9.04 (16.42) |
| Drink alcohol daily (%) | 20.48 | 16.5 | 10.21 |
| Monthly Units | 56.73 (72.52) | 57.16 (83.88) | 51.65 (76.4) |
| Sleep Duration (h) | 7.05 (1.03) | 6.91 (1.26) | 6.67 (1.52) |
| Morning Chronotype (%) | 23.33 | 24.37 | 19.24 |
| Evening Chronotype (%) | 8.02 | 8.7 | 16.9 |
| **Ethnicity (%):** |  |  |  |
| White British | 88.47 | 81.84 | 80.99 |
| White Other | 6.45 | 7.05 | 6.01 |
| Mixed | 0.65 | 0.93 | 0.87 |
| Asian | 1.72 | 3.69 | 3.39 |
| Black | 1.4 | 3.64 | 5.47 |
| Chinese | 0.34 | 0.47 | 0.67 |
| Other | 6.45 | 7.05 | 6.01 |
| Weekly work hours | 34.72 (12.6) | 37.62 (12.93) | 40.01 (13.16) |
|  | Day workers | Irregular shift work | Permanent night shift work |
| Single Occupancy (%) | 15.64 | 18.75 | 18.42 |
| Townsend Index | -2.24 (-3.7 to 0.19) | -1.28 (-3.18 to 1.71) | -1.04 (-3.02 to 2.07) |
| Maternal Smoking (%) | 26.59 | 29.03 | 30.75 |
| Birth Weight (kg) | 3.33 (0.63) | 3.33 (0.68) | 3.31 (0.71) |
| High Cholesterol (%) | 7.88 | 8.55 | 9.27 |
| All Diabetes (%) | 3.23 | 4.42 | 4.68 |
| Type II Diabetes (%) | 0.41 | 0.59 | 0.59 |
| All Hypertension (%) | 20.33 | 22.16 | 23.31 |
| Essential Hypertension (%) | 0.28 | 0.33 | 0.35 |
| Depression (%) | 4.62 | 5.07 | 4.79 |
| Cardiovascular Disease (%) | 3.82 | 4.31 | 4.09 |
| DSI | -2.65 (1.39) | -2.41 (1.42) | -2.26 (1.39) |
| Probability of NAFLD (from DSI score) | 0.12 (0.14) | 0.14 (0.15) | 0.15 (0.16) |

Data are mean (SD) or percentages.
DSI (Dallas Steatosis Index) and Fib 4 (Fibrosis 4) are defined in Appendix A.

**Supplementary Table 2a:** Adjusted odds (95% CI) of having high probability of NAFLD (p > 0.6, calculated from the DSI) by shift work schedule (N = 240,589)

|  | Current work schedule |  |  |
| --- | --- | --- | --- |
|  | Day workers | Irregular shift work | Always |
| Total cases (% of total sample size) | 2,816 (1.42%) | 761 (2.13%) | 142 (2.36%) |
| Total sample size | 198,854 | 35,712 | 6,023 |
| Model 1: Age, Sex, Ethnicity and TDI adjusted. | 1 | 1.43 (1.32-1.55) | 1.57 (1.32-1.86) |
| Model 2: Model 1 covariates + Sleep Duration, Alcohol, Smoking and Length of working week. | 1 | 1.29 (1.18-1.4) | 1.26 (1.06-1.5) |
| Model 3: Model 2 covariates + BMI | 1 | 1.12 (1.03-1.22) | 1.08 (0.9-1.29) |

**Supplementary Table 2b:** Adjusted means (standard error) of DSI by shift work schedule (N = 240,589)

|  | Current work schedule |  |  |
| --- | --- | --- | --- |
|  | Day workers | Irregular shift work | Always |
| Total sample size | 198,854 | 35,712 | 6,023 |
| Model 1: Age, Sex, Ethnicity and TDI adjusted. | ref | 0.19 (0.01)*** | 0.32 (0.02)*** |
| Model 2: Model 1 covariates + Sleep Duration, Alcohol, Smoking and Length of working week. | ref | 0.15 (0.01)*** | 0.22 (0.02)*** |
| Model 3: Model 2 covariates + BMI | ref | 0.03 (0.01)*** | 0.05 (0.01)*** |

Statistical significance using Student's t-test. \* indicates $p < 0.05$ , \*\* $p < 0.01$ and \*\*\* $p < 0.001$ . "ref" signifies reference group for calculation of adjusted means.

**Supplementary Table 3:** Adjusted odds (95% CI) of NAFLD or NASH ICD10 code by shift work schedule (N = 280,116)

|  | Current work schedule |  |  |
| --- | --- | --- | --- |
|  | Day workers | Irregular shift work | Always |
| Total cases (% of total sample size) | 174 (0.08%) | 45 (0.11%) | 7 (0.1%) |
| Total sample size | 231,747 | 41,375 | 6,994 |
| Model 1: Age, Sex, Ethnicity and TDI adjusted. | 1 | 1.37 (0.98-1.91) | 1.23 (0.58-2.63) |
| Model 2: Model 1 covariates + Sleep Duration, Alcohol, Smoking and Length of working week. | 1 | 1.24 (0.89-1.74) | 1.01 (0.47-2.18) |
| Model 3: Model 2 covariates + BMI | 1 | 1.16 (0.83-1.62) | 0.93 (0.43-1.99) |

**Supplementary Table 4a:** Adjusted odds (95% CI) of high proton density fat fraction (> 5.5%) by shift work schedule (N = 6,419)

|  | Current work schedule |  |  |
| --- | --- | --- | --- |
|  | Day workers | Irregular shift work | Always |
| Total cases (% of total sample size) | 1,145 (20.7%) | 194 (25.1%) | 24 (22.0%) |
| Total sample size | 5,537 | 773 | 109 |
| Model 1: Age, Sex, Ethnicity and TDI adjusted. | 1 | 1.26 (1.06-1.51) | 1.02 (0.64-1.62) |
| Model 2: Model 1 covariates + Sleep Duration, Alcohol, Smoking and Length of working week. | 1 | 1.18 (0.98-1.41) | 0.9 (0.56-1.44) |
| Model 3: Model 2 covariates + BMI | 1 | 1.11 (0.92-1.35) | 0.9 (0.55-1.47) |

**Supplementary Table 4b:** Adjusted means (standard error) of proton density fat fraction by shift work schedule (N = 6,419).

|  | Current work schedule |  |  |
| --- | --- | --- | --- |
|  | Day workers | Irregular shift work | Always |
| Total sample size | 5,537 | 773 | 109 |
| Model 1: Age, Sex, Ethnicity and TDI adjusted. | ref | 0.57 (0.18)** | -0.02 (0.45) |
| Model 2: Model 1 covariates + Sleep Duration, Alcohol, Smoking and Length of working week. | ref | 0.43 (0.18)* | -0.27 (0.45) |
| Model 3: Model 2 covariates + BMI | ref | 0.3 (0.17) | -0.25 (0.42) |

Statistical significance using Student's t-test. \* indicates $p < 0.05$ , \*\* $p < 0.01$ and \*\*\* $p < 0.001$ . "ref" signifies reference group for calculation of adjusted means.

**Supplementary Table 5a:** Adjusted odds (95% CI) of having high probability of NAFLD (p > 0.6, calculated from the DSI) by chronotype (N = 377,560)

|  | Chronotype |  |  |
| --- | --- | --- | --- |
|  | Intermediate chronotype | Definitely a morning person | Definitely an evening person |
| Total cases (% of total sample size) | 4,174 (1.73%) | 2,011 (1.97%) | 924 (2.72%) |
| Total sample size | 241,398 | 102,172 | 33,990 |
| Model 1: Age, Sex, Ethnicity and TDI adjusted. | 1 | 1.09 (1.03-1.15) | 1.55 (1.44-1.66) |
| Model 2: Model 1 covariates + Sleep Duration, Alcohol, Smoking and Length of working week. | 1 | 1.03 (0.98-1.09) | 1.45 (1.35-1.56) |
| Model 3: Model 2 covariates + BMI | 1 | 0.99 (0.94-1.05) | 1.29 (1.19-1.39) |

**Supplementary Table 5b:** Adjusted means (standard error) of DSI by chronotype (N = 377,560)

|  | Chronotype |  |  |
| --- | --- | --- | --- |
|  | Intermediate chronotype | Definitely a morning person | Definitely an evening person |
| Total sample size | 241,398 | 102,172 | 33,990 |
| Model 1: Age, Sex, Ethnicity and TDI adjusted. | ref | 0.06 (0.01)*** | 0.16 (0.01)*** |
| Model 2: Model 1 covariates + Sleep Duration, Alcohol, Smoking and Length of working week. | ref | 0.03 (0.01)*** | 0.14 (0.01)*** |
| Model 3: Model 2 covariates + BMI | ref | 0.00 (0.00) | 0.05 (0.01)*** |

Statistical significance using Student's t-test. \* indicates $p < 0.05$ , \*\* $p < 0.01$ and \*\*\* $p < 0.001$ . "ref" signifies reference group for calculation of adjusted means.

**Supplementary Table 6:** Adjusted odds (95% CI) of NAFLD or NASH ICD10 code by chronotype (N = 435,830)

|  | Chronotype |  |  |
| --- | --- | --- | --- |
|  | Intermediate chronotype | Definitely a morning person | Definitely an evening person |
| Total cases (% of total sample size) | 305 (0.11%) | 142 (0.12%) | 55 (0.14%) |
| Total sample size | 278,627 | 118,064 | 39,139 |
| Model 1: Age, Sex, Ethnicity and TDI adjusted. | 1 | 1.04 (0.85-1.27) | 1.24 (0.93-1.65) |
| Model 2: Model 1 covariates + Sleep Duration, Alcohol, Smoking and Length of working week. | 1 | 0.97 (0.8-1.19) | 1.14 (0.85-1.52) |
| Model 3: Model 2 covariates + BMI | 1 | 0.96 (0.78-1.17) | 1.08 (0.81-1.44) |

**Supplementary Table 7a:** Adjusted odds (95% CI) of high proton density fat fraction (> 5.5%) by chronotype (N = 8,651)

|  | Chronotype |  |  |
| --- | --- | --- | --- |
|  | Intermediate chronotype | Definitely a morning person | Definitely an evening person |
| Total cases (% of total sample size) | 1,181 (20.6%) | 454 (21.9%) | 200 (23.7%) |
| Total sample size | 5,737 | 2,070 | 844 |
| Model 1: Age, Sex, Ethnicity and TDI adjusted. | 1 | 1.08 (0.96-1.23) | 1.16 (0.98-1.38) |
| Model 2: Model 1 covariates + Sleep Duration, Alcohol, Smoking and Length of working week. | 1 | 1.07 (0.95-1.22) | 1.1 (0.92-1.31) |
| Model 3: Model 2 covariates + BMI | 1 | 1.04 (0.91-1.19) | 1 (0.83-1.20) |

**Supplementary Table 7b:** Adjusted means (standard error) of proton density fat fraction by chronotype (N = 8,651).

|  | Chronotype |  |  |
| --- | --- | --- | --- |
|  | Intermediate chronotype | Definitely a morning person | Definitely an evening person |
| Total sample size | 5,737 | 2,070 | 844 |
| Model 1: Age, Sex, Ethnicity and TDI adjusted. | ref | 0.21 (0.15) | 0.44 (0.21)* |
| Model 2: Model 1 covariates + Sleep Duration, Alcohol, Smoking and Length of working week. | ref | 0.21 (0.15) | 0.32 (0.21) |
| Model 3: Model 2 covariates + BMI | ref | 0.16 (0.14) | 0.13 (0.2) |

Statistical significance using Student's t-test. \* indicates $p < 0.05$ , \*\* $p < 0.01$ and \*\*\* $p < 0.001$ . "ref" signifies reference group for calculation of adjusted means.

**Supplementary Table 8:**
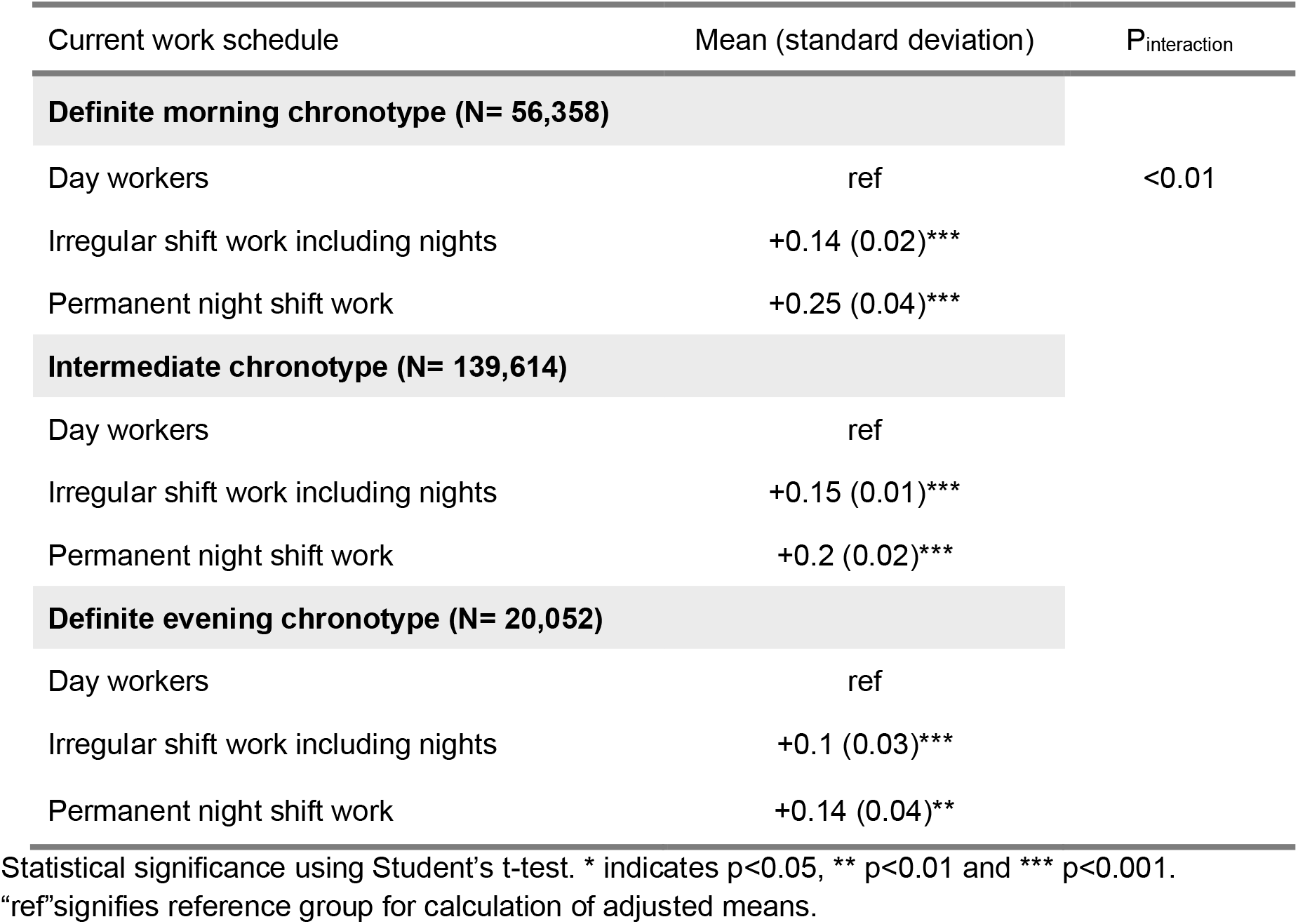
Change in adjusted means (standard deviation) of DSI by shift work stratified by chronotype (compared to day workers)

## Appendix A

The Dallas Steatosis Index (DSI) was defined as a LOGIT equation as derived in McHenry et al (2020):

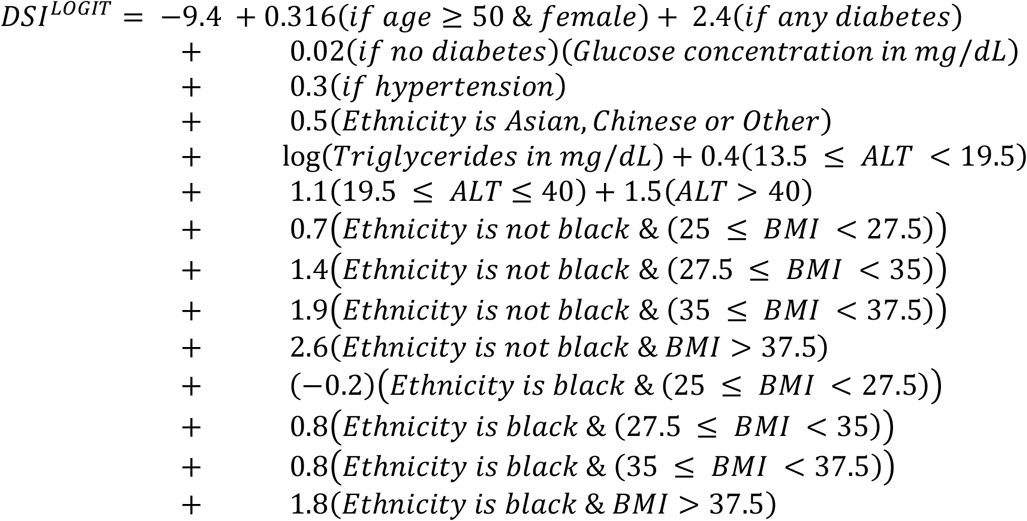

We defined the probability of NAFLD using the following formula and defined high-risk as a probability >0.6:

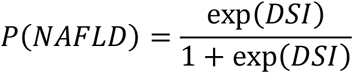

## References

[1] Younossi ZM, Koenig AB, Abdelatif D, Fazel Y, Henry L, Wymer M. Global epidemiology of nonalcoholic fatty liver disease—Meta-analytic assessment of prevalence, incidence, and outcomes. Hepatology 2016;64:73–84. https://doi.org/10.1002/hep.28431.

[2] Marchesini G, Day CP, Dufour JF, Canbay A, Nobili V, Ratziu V, et al. EASL-EASD-EASO Clinical Practice Guidelines for the Management of Non-Alcoholic Fatty Liver Disease. Obes Facts 2016;9:65. https://doi.org/10.1159/000443344.

[3] Bass J, Takahashi JS. Circadian integration of metabolism and energetics. Science 2010;330:1349–54. https://doi.org/10.1126/science.1195027.

[4] Allada R, Bass J. Circadian Mechanisms in Medicine. N Engl J Med 2021;384:550–61. https://doi.org/10.1056/nejmra1802337.

[5] Zhang S, Wang Y, Wang Z, Wang H, Xue C, Li Q, et al. Rotating night shift work and non-Alcoholic fatty liver disease among steelworkers in China: A cross-sectional survey. Occup Environ Med 2020;77:333–9. https://doi.org/10.1136/oemed-2019-106220.

[6] Wang F, Zhang L, Wu S, Li W, Sun M, Feng W, et al. Night shift work and abnormal liver function: Is non-alcohol fatty liver a necessary mediator? Occup Environ Med 2019;76:83–9. https://doi.org/10.1136/oemed-2018-105273.

[7] Balakrishnan M, El-Serag HB, Kanwal F, Thrift AP. Shiftwork Is Not Associated with Increased Risk of NAFLD: Findings from the National Health and Nutrition Examination Survey. Dig Dis Sci 2017;62:526–33. https://doi.org/10.1007/s10620-016-4401-1.

[8] McHenry S, Park Y, Davidson NO. Validation of the Dallas Steatosis Index to Predict Nonalcoholic Fatty Liver Disease in the UK Biobank Population. Clin Gastroenterol Hepatol 2021. https://doi.org/10.1016/J.CGH.2021.05.035.

[9] Wilman HR, Kelly M, Garratt S, Matthews PM, Milanesi M, Herlihy A, et al. Characterisation of liver fat in the UK Biobank cohort. PLoS One 2017;12:e0172921. https://doi.org/10.1371/journal.pone.0172921.

[10] Kitamura S, Hida A, Aritake S, Higuchi S, Enomoto M, Kato M, et al. Validity of the Japanese version of the Munich ChronoType Questionnaire. Chronobiol Int 2014;31:845–50. https://doi.org/10.3109/07420528.2014.914035.

[11] Megdal SP, Schernhammer ES. Correlates for poor sleepers in a Los Angeles high school. Sleep Med 2007;9:60–3. https://doi.org/10.1016/J.SLEEP.2007.01.012.

[12] Taillard J, Philip P, Chastang JF, Bioulac B. Validation of Horne and Ostberg morningness-eveningness questionnaire in a middle-aged population of French workers. J Biol Rhythms 2004;19:76–86. https://doi.org/10.1177/0748730403259849.

[13] Wilman HR, Kelly M, Garratt S, Matthews PM, Milanesi M, Herlihy A, et al. Characterisation of liver fat in the UK Biobank cohort. PLoS One 2017;12. https://doi.org/10.1371/journal.pone.0172921.

[14] Maidstone RJ, Turner J, Vetter C, Dashti HS, Saxena R, Scheer Fajl, et al. Night shift work is associated with an increased risk of asthma. Thorax 2021;76:53–60. https://doi.org/10.1136/thoraxjnl-2020-215218.

[15] Roenneberg T, Winnebeck EC, Klerman EB. Daylight Saving Time and Artificial Time Zones – A Battle Between Biological and Social Times. Front Physiol 2019;10:944. https://doi.org/10.3389/fphys.2019.00944.

[16] Ge X, Zheng L, Wang M, Du Y, Jiang J. Prevalence trends in non-alcoholic fatty liver disease at the global, regional and national levels, 1990-2017: a population-based observational study. BMJ Open 2020;10:e036663. https://doi.org/10.1136/bmjopen-2019-036663.

[17] Estes C, Razavi H, Loomba R, Younossi Z, Sanyal AJ. Modeling the epidemic of nonalcoholic fatty liver disease demonstrates an exponential increase in burden of disease. Hepatology 2018;67:123–33. https://doi.org/10.1002/hep.29466.

[18] Kettner NM, Voicu H, Finegold MJ, Coarfa C, Sreekumar A, Putluri N, et al. Circadian Homeostasis of Liver Metabolism Suppresses Hepatocarcinogenesis. Cancer Cell 2016;30:909–24. https://doi.org/10.1016/j.ccell.2016.10.007.

[19] Mukherji A, Kobiita A, Damara M, Misra N, Meziane H, Champy MF, et al. Shifting eating to the circadian rest phase misaligns the peripheral clocks with the master SCN clock and leads to a metabolic syndrome. Proc Natl Acad Sci U S A 2015;112:E6691–8. https://doi.org/10.1073/pnas.1519807112.

[20] Daghlas I, Richmond RC, Lane JM, Dashti HS, Ollila HM, Schernhammer ES, et al. Selection into shift work is influenced by educational attainment and body mass index: A Mendelian randomization study in the UK Biobank. Int J Epidemiol 2021;50:1229–40. https://doi.org/10.1093/ije/dyab031.

[21] Liu Q, Shi J, Duan P, Liu B, Li T, Wang C, et al. Is shift work associated with a higher risk of overweight or obesity? A systematic review of observational studies with meta-analysis. Int J Epidemiol 2018;47:1956–71. https://doi.org/10.1093/ije/dyy079.

[22] Ganz M, Szabo G. Immune and inflammatory pathways in NASH. Hepatol Int 2013;7:S771–81. https://doi.org/10.1007/s12072-013-9468-6.

[23] Seki E, Schwabe RF. Hepatic inflammation and fibrosis: Functional links and key pathways. Hepatology 2015;61:1066–79. https://doi.org/10.1002/hep.27332.

[24] Heymann F, Tacke F. Immunology in the liver — from homeostasis to disease. Nat Publ Gr 2016. https://doi.org/10.1038/nrgastro.2015.200.

[25] Samuel VT, Shulman GI. Nonalcoholic Fatty Liver Disease as a Nexus of Metabolic and Hepatic Diseases. Cell Metab 2018;27:22–41. https://doi.org/10.1016/j.cmet.2017.08.002.

[26] Haas JT, Francque S, Staels B. Pathophysiology and Mechanisms of Nonalcoholic Fatty Liver Disease. Annu Rev Physiol 2016;78:181–205. https://doi.org/10.1146/annurev-physiol-021115-105331.

[27] Baxter M, Ray DW. Circadian rhythms in innate immunity and stress responses. Immunology 2019. https://doi.org/10.1111/imm.13166.

[28] Marjot T, Ray DW, Tomlinson JW. Is it time for chronopharmacology in NASH? J Hepatol 2022;0. https://doi.org/10.1016/j.jhep.2021.12.039.

[29] Caratti G, Iqbal M, Hunter L, Kim D, Wang P, Vonslow RM, et al. REVERBa couples the circadian clock to hepatic glucocorticoid action. J Clin Investig 2018;128:4454–71. https://doi.org/10.1172/JCI96138.

